# Risk stratification of patients admitted to hospital with covid-19 using the ISARIC WHO Clinical Characterisation Protocol: development and validation of the 4C Mortality Score

**DOI:** 10.1101/2020.07.30.20165464

**Authors:** Stephen R Knight, Antonia Ho, Riinu Pius, Iain Buchan, Gail Carson, Thomas M Drake, Jake Dunning, Cameron J Fairfield, Carrol Gamble, Christopher A Green, Rishi Gupta, Sophie Halpin, Hayley E Hardwick, Karl A Holden, Peter W Horby, Clare Jackson, Kenneth A Mclean, Laura Merson, Jonathan S Nguyen-Van-Tam, Lisa Norman, Mahdad Noursadeghi, Piero L Olliaro, Mark G Pritchard, Clark D Russell, Catherine A Shaw, Aziz Sheikh, Tom Solomon, Cathie Sudlow, Olivia V Swann, Lance CW Turtle, Peter JM Openshaw, J Kenneth Baillie, Malcolm G Semple, Annemarie B Docherty, Ewen M Harrison, on behalf of the ISARIC4C investigators

## Abstract

**Objectives:** To develop and validate a pragmatic risk score to predict mortality for patients admitted to hospital with covid-19.

**Design:** Prospective observational cohort study: ISARIC WHO CCP-UK study (ISARIC Coronavirus Clinical Characterisation Consortium [4C]). Model training was performed on a cohort of patients recruited between 6 February and 20 May 2020, with validation conducted on a second cohort of patients recruited between 21 May and 29 June 2020.

**Setting:** 260 hospitals across England, Scotland, and Wales.

**Participants:** Adult patients (≥18 years) admitted to hospital with covid-19 admitted at least four weeks before final data extraction.

**Main outcome measures:** In-hospital mortality.

**Results:** There were 34 692 patients included in the derivation dataset (mortality rate 31.7%) and 22 454 in the validation dataset (mortality 31.5%). The final 4C Mortality Score included eight variables readily available at initial hospital assessment: age, sex, number of comorbidities, respiratory rate, peripheral oxygen saturation, level of consciousness, urea, and C-reactive protein (score range 0-21 points). The 4C risk stratification score demonstrated high discrimination for mortality (derivation cohort: AUROC 0.79; 95% CI 0.78 − 0.79; validation cohort 0.78, 0.77-0.79) with excellent calibration (slope = 1.0). Patients with a score ≥15 (n = 2310, 17.4%) had a 67% mortality (i.e., positive predictive value 67%) compared with 1.0% mortality for those with a score ≤3 (n = 918, 7%; negative predictive value 99%). Discriminatory performance was higher than 15 pre-existing risk stratification scores (AUROC range 0.60-0.76), with scores developed in other covid-19 cohorts often performing poorly (range 0.63-0.73).

**Conclusions:** We have developed and validated an easy-to-use risk stratification score based on commonly available parameters at hospital presentation. This outperformed existing scores, demonstrated utility to directly inform clinical decision making, and can be used to stratify inpatients with covid-19 into different management groups. The 4C Mortality Score may help clinicians identify patients with covid-19 at high risk of dying during current and subsequent waves of the pandemic.

**Study registration:** ISRCTN66726260

## Introduction

Disease resulting from infection with severe acute respiratory syndrome coronavirus 2 (SARS-CoV-2), has a high mortality rate with deaths predominantly due to respiratory failure.^1^ As of 30^th^ July 2020, there are over 17 million confirmed cases worldwide and at least 660 000 deaths.^2,3^ As hospitals around the world are faced with an influx of patients with covid-19, there is an urgent need for a pragmatic risk stratification tool that will allow the early identification of patients infected with SARS-CoV-2 who are at the highest risk of death, to guide management and optimise resource allocation.

Prognostic scores attempt to transform complex clinical pictures into tangible numerical values. Prognostication is more difficult when dealing with a severe pandemic illness such as covid-19, as strain on healthcare resources and rapidly evolving treatments alter the risk of death over time. Early information has suggested that the clinical course of a patient with covid-19 is different from that of pneumonia, seasonal influenza or sepsis.^4^ The majority of patients with severe covid-19 have developed a clinical picture characterised by pneumonitis, profound hypoxia, and systemic inflammation affecting multiple organs.^1^

A recent review identified many prognostic scores used for covid-19,^5^ which varied in their setting, predicted outcome measure, and the clinical parameters included. The large number of risk stratification tools reflects difficulties in their application, with most scores demonstrating moderate performance at best and no benefit to clinical decision-making.^6,7^ It has been found that many novel covid-19 prognostic scores have a high risk of bias, which may reflect development in small cohorts, and many have been published without clear details of model derivation and testing.^5^ To our knowledge, a risk stratification tool is yet to be developed and validated within a large national cohort of hospitalised patients with covid-19.

Our aim was to develop and validate a pragmatic, clinically relevant risk stratification score using routinely available clinical information at hospital presentation to predict in-hospital mortality in hospitalised covid-19 patients and then compare this with existing prognostic models.

## Methods

### Study design and setting

The International Severe Acute Respiratory and emerging Infections Consortium (ISARIC) WHO Clinical Characterisation Protocol UK (CCP-UK) study is an ongoing prospective cohort study in 260 acute care hospitals across England, Scotland, and Wales (National Institute for Health Research Clinical Research Network Central Portfolio Management System ID: 14152) performed by the ISARIC Covid-19 Clinical Characterisation Consortium (ISARIC-4C). The protocol and further study details are available online.^8^ Model development and reporting followed the Transparent Reporting of a multivariate prediction mode for Individual Prediction or Diagnosis (TRIPOD) guidelines.^9^ The study was performed according to a pre-defined protocol (Appendix 1).

### Participants

Patients aged ≥18 years old with a completed index admission to one of 260 hospitals in England, Scotland, or Wales were included.^8^ Reverse transcriptase-PCR was the only mode of testing available during the period of study. The decision to test was at the discretion of the clinician attending the patient, and not defined by protocol. The enrolment criterion “high likelihood of infection” reflected that a preparedness protocol cannot assume a diagnostic test will be available for an emergent pathogen. In this activation, site training emphasised importance of only recruiting proven cases.

### Data collection

Demographic, clinical and outcomes data were collected using a pre-specified case report form. Comorbidities were defined according to a modified Charlson Comorbidity Index^10^; those collected were: chronic cardiac disease; chronic respiratory disease (excluding asthma); chronic renal disease (estimated glomerular filtration rate ≤30); mild-to-severe liver disease; dementia; chronic neurological conditions; connective tissue disease; diabetes mellitus (diet, tablet or insulin-controlled); HIV/AIDS, and malignancy. These were selected *a priori* by a global consortium to provide rapid, coordinated clinical investigation of patients presenting with any severe or potentially severe acute infection of public interest and enabled standardisation.

Clinician-defined obesity was also included as a comorbidity due its likely association with adverse outcomes in patients with covid-19.^11,12^ Patients with missing data on all comorbidities were assumed to have no comorbidities. The clinical information used to calculate prognostic scores was taken from the day of admission to hospital.^13^ No generally accepted approaches exist to estimate sample size requirements for derivation and validation studies of risk prediction models. We used all available data to maximise the power and generalisability of our results. Model reliability was enhanced by our use of a validation cohort and sensitivity analyses

### Outcomes

The primary outcome was in-hospital mortality. This outcome was selected due to the importance of the early identification of patients likely to develop severe illness from SARS-CoV-2 infection (a ‘rule in’ test). We chose *a priori* to restrict analysis of outcomes to patients who were admitted more than four weeks before final data extraction (29^th^ June 2020) to enable most patients to complete their hospital admission.

## Independent predictor variables

A reduced set of potential predictor variables was selected *a priori* including patient demographic information, common clinical investigations, and parameters consistently identified as clinically important in covid-19 cohorts following methodology described by Wynants et al.^5^ Candidate predictor variables were selected based on three common criteria:^14^ (1) patient and clinical variables known to influence outcome in pneumonia and flu-like illness; (2) clinical biomarkers previously identified within the literature as potential predictors in covid-19 patients; and (3) values were available for at least two-thirds of patients within the derivation cohort.

With the overall aim to develop an easy-to-use risk stratification score, an *a priori* decision was made to include an overall comorbidity count for each patient within model development giving each comorbidity equal weight, rather than individual comorbidities. Recent evidence suggests an additive effect of comorbidity in covid-19 patients, with increasing number of comorbidities associated with poorer outcomes.^15^

### Model development

Models were trained using all available data up to 20^th^ May 2020. The primary intention was to create a pragmatic model for bedside use not requiring complex equations, online calculators, or mobile applications. An *a priori* decision was therefore made to categorise continuous variables in the final prognostic score.

A three-stage model building process was used (Figure 1). Firstly, generalised additive models (GAMs) were built incorporating continuous smoothed predictors (thin-plate splines) in combination with categorical predictors as linear components. A criterion-based approach to variable selection was taken based on the deviance explained, the unbiased risk estimator, and the area under the receiver operating characteristic curve (AUROC) (see Appendix 2).

**Figure 1.**
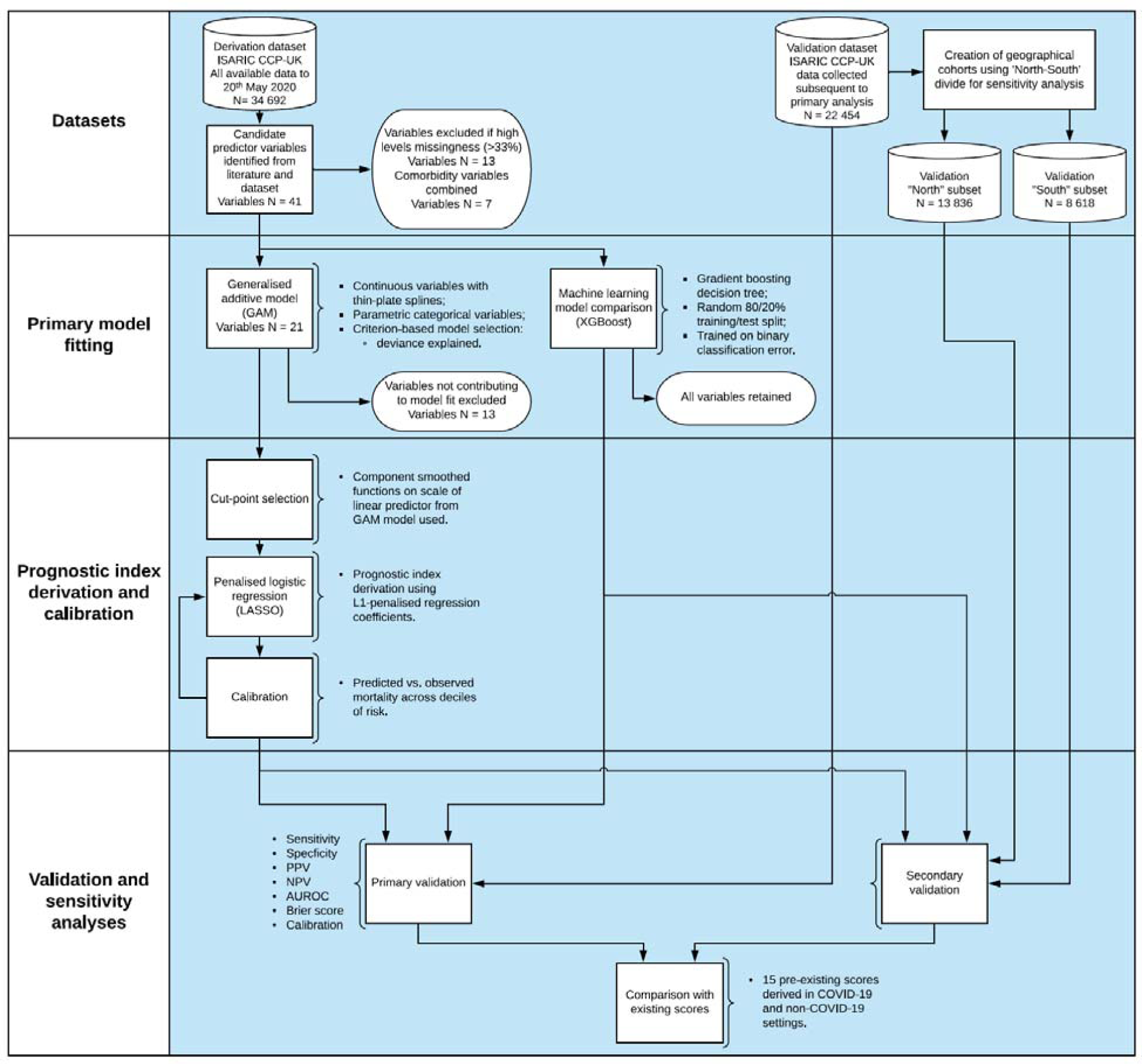
Model derivation and validation workflow.

Secondly, plots of component smoothed continuous predictors were visually inspected for linearity and optimal cut-points were selected using the methods of Barrio *et* al.^16^

Lastly, final models using categorised variables were specified using least absolute shrinkage and selection operator (LASSO) logistic regression. L1-penalised coefficients were derived using 10-fold cross-validation to select the value of lambda (minimised cross-validated sum of squared residuals). Shrunk coefficients were converted to a prognostic index with appropriate scaling to create the pragmatic “4C” Mortality Score (where 4C stands for Coronavirus Clinical Characterisation Consortium).

Machine learning approaches were used in parallel for comparison of predictive performance. Given issues with interpretability, this was intended to provide a “best-in-class” comparison of predictive performance when accounting for any complex underlying interactions. Extreme gradient boosting trees were used (XGBoost). All candidate predictor variables identified were included within the model, with the exception of those with high missing values (>33%). Individual major comorbidity variables, defined as chronic cardiac disease; chronic respiratory disease (excluding asthma); chronic renal disease (estimated glomerular filtration rate ≤30 mL/min/1.73m^2^); moderate-to-severe liver failure (presence of portal hypertension); diabetes mellitus (diet, tablet or insulin-controlled) and solid malignancy, together with obesity, were retained within the model to determine whether their inclusion enhanced predictive performance. An 80%/20% random split of the derivation dataset was used to define train/test sets. The validation datasets were held back and not used in the training process. A mortality label and design matrix of centred/standardised continuous and categorical variables including all candidate variables was used to train gradient boosted trees minimising the binary classification error rate (defined as number wrong cases / number all cases). Hyperparameters were tuned including the learning rate and maximum tree depth to maximise the AUROC in the test set.

Discrimination was assessed for all above models (4C and XGBoost model) using the AUROC in the derivation cohort, with 95% confidence intervals (CI) calculated using bootstrapping resampling (2000 samples). An AUROC value of 0.5 indicates no predictive ability, 0.8 is considered good, and 1.0 is perfect.^17^ Overall goodness-of-fit was assessed with the Brier score,^18^ a measure to quantify how close predictions are to the truth ranging between 0 and 1, where smaller values indicate superior model performance. We plotted model calibration curves to examine agreement between predicted and observed risk across deciles of mortality risk to ascertain the presence of over- or under-prediction. Risk cut-off values were defined by the total point score for an individual which represented a low (<2% mortality rate), intermediate (2-14.9%) or high-risk (≥15%) groups, similar to commonly used pneumonia risk stratification scores.^19,20^

Sensitivity analyses of missing values in potential candidate variables were performed using multiple imputation by chained equations, under the missing at random assumption. Ten sets, each with 10 iterations, were imputed using available explanatory variables for both cohorts (derivation and validation). The outcome variable was included as a predictor in the derivation but not validation dataset. Model derivation was explored in imputed datasets. All models developed in the complete case derivation dataset were tested in the imputed validation dataset, with Rubin’s rules^21^ used to combine model parameter estimates.

### Model validation

Patients entered subsequently into the ISARIC WHO CCP-UK study after 20^th^ May 2020 were included in a separate validation cohort (Figure 1). We determined discrimination, calibration, and performance across a range of clinically relevant metrics. To avoid bias in the assessment of outcomes, patients who admitted within four weeks prior to data extraction on 29^th^ June 2020 were excluded.

A sensitivity analysis was also performed, with stratification of the validation cohort by geographical location. This geographical categorisation was selected based on well-described economic and health inequalities between the north and south of the UK.^22,23^ Recent analysis has demonstrated the impact of deprivation on risk of dying with covid-19.^24^ As a result, population differences between regions may change the discriminatory performance of risk stratification scores. Two geographical cohorts were created, based on north-south geographical locations across the United Kingdom as defined by Hacking et al.^22^ A further sensitivity analysis was performed to determine model performance in ethnic minority groups, given reported differences in covid-19 outcomes.^25^

All tests were two-tailed and p values <0.05 were considered statistically significant. We used R (version 3.6.3) with the *finalfit, glmnet, pROC, recipes, xgboost, rmda*, and *tidyverse* packages for all statistical analysis.

### Comparison with existing risk stratification scores

All derived models in the derivation dataset were compared within the validation cohort with existing scores. Model performance was assessed using the AUROC statistic, sensitivity, specificity, positive predictive value (PPV), and negative predictive value (NPV). Existing risk stratification scores were identified through a systematic literature search of EMBASE, WHO Medicus, and Google Scholar databases. We used the search terms “pneumonia”, “sepsis”, “influenza”, “COVID-19”, “SARS-CoV-2”, “coronavirus” combined with “score”‘ and “prognosis”. We applied no language or date restrictions. The last search was performed on 1^st^ July 2020. Risk stratification tools were included whose variables were available within the database and had accessible methodology for calculation.

Performance characteristics were calculated according to original publications, and score cut-offs for adverse outcomes were selected based on the most commonly used criteria identified during the literature search. Cut-offs were the score value for which the patient was considered at low- or high-risk of adverse outcome, as defined by study authors. Patients with one or more missing input variables were omitted for that particular score.

A decision curve analysis (DCA) was also performed.^26^ Briefly, assessment of the adequacy clinical prediction models can be extended by determining clinical utility. Using DCA, a clinical judgment of the relative value of benefits (treating a true positive) and harms (treating a false positive) associated with a clinical prediction tool can be made. The standardised net benefit was plotted against the threshold probability for considering a patient “high risk” for age alone and the best discriminating models applicable to >50% of patients in the validation cohort.

### Patient and public involvement

This was an urgent public health research study in response to a Public Health Emergency of International Concern. Patients or the public were not involved in the design, conduct, or reporting of this rapid response research.

## Results

In the derivation cohort, we collected data from 34 692 patients between 6^th^ February 2020 and 20^th^ May 2020. The overall mortality was 31.7% (10 998 patients). The median age of patients in the cohort was 74 (interquartile range (IQR) 59-83) years. 58.3% (20 184) were male and 75.3% (26 135) patients had at least one comorbidity. Demographic and clinical characteristics for the derivation and validation datasets are shown in Table 1.

**Table 1.** Demographic and clinical characteristics for derivation and validation cohorts for patients hospitalized with covid-19.

|  |  | Derivation cohort<br>(n = 34 692) | Validation cohort<br>(n = 22 454) |
| --- | --- | --- | --- |
| Mortality (%) |  | 10 998 (31.7) | 6428 (28.6) |
| Age on admission (years) | <50 | 4397 (12.7) | 2825 (12.6) |
|  | 50-59 | 4603 (13.3) | 2630 (11.7) |
|  | 60-69 | 5563 (16.0) | 3155 (14.1) |
|  | 70-79 | 7986 (23.0) | 4971 (22.1) |
|  | ≥80 | 12 143 (35.0) | 8873 (39.5) |
| Sex at Birth | Male | 20 184 (58.3) | 12 202 (54.4) |
|  | Female | 14 411 (41.7) | 10 211 (45.6) |
| Ethnicity | White | 25 680 (83.2) | 16 837 (85.0) |
|  | South Asian | 1423 (4.6) | 786 (4.0) |
|  | East Asian | 269 (0.9) | 138 (0.7) |
|  | Black | 1108 (3.6) | 765 (3.9) |
|  | Other Ethnic Minority | 2387 (7.7) | 1275 (6.4) |
| Chronic cardiac disease |  | 10 192 (32.0) | 6853 (33.9) |
| Chronic kidney disease |  | 5451 (17.3) | 3682 (18.4) |
| Malignant neoplasm |  | 3201 (10.2) | 2144 (10.8) |
| Moderate or severe liver disease |  | 581 (1.9) | 427 (2.2) |
| Clinician-reported obesity |  | 3250 (11.3) | 2127 (11.9) |
| Chronic pulmonary disease (not asthma) |  | 5684 (17.9) | 3671 (18.3) |
| Diabetes |  | 8245 (26.3) | 4210 (22.0) |
| Number of comorbidities | 0 | 8557 (24.7) | 5524 (24.6) |
|  | 1 | 9633 (27.8) | 6045 (26.9) |
|  | ≥2 | 16 502 (47.6) | 10 885 (48.5) |
| Respiratory Rate |  | 22.0 (8.0) | 20.0 (8.0) |
| Peripheral oxygen saturation (%) |  | 94.0 (6.0) | 94.0 (5.0) |
| Systolic blood pressure (mmHg) |  | 125.0 (33.0) | 129.0 (33.0) |
| Diastolic blood pressure (mmHg) |  | 70.0 (19.0) | 73.0 (20.0) |
| Temperature (°C) |  | 37.3 (1.6) | 37.1 (1.5) |
| Heart Rate (bpm) |  | 90.0 (27.0) | 90.0 (28.0) |
| Glasgow Coma Score |  | 15.0 (0.0) | 15.0 (0.0) |
| pH |  | 7.4 (0.1) | 7.4 (0.1) |
| Bicarbonate (mmol/L) |  | 24.5 (4.5) | 24.4 (5.0) |
| Infiltrates on chest radiograph |  | 13 984 (62.9) | 8244 (61.1) |
| Haemoglobin (g/L) |  | 130.0 (29.0) | 127.0 (31.0) |

|  | Derivation cohort<br>(n = 34 692) | Validation cohort<br>(n = 22 454) |
| --- | --- | --- |
| White cell count ($10^9$/L) | 7.4 (5.0) | 7.6 (5.3) |
| Neutrophil count ($10^9$/L) | 5.6 (4.6) | 5.8 (4.9) |
| Lymphocyte count ($10^9$/L) | 0.9 (0.6) | 0.9 (0.7) |
| Haematocrit (%) | 35.0 (40.6) | 25.2 (38.6) |
| Platelet Count ($10^9$/L) | 215.0 (120.0) | 223.0 (126.0) |
| Prothrombin (seconds) | 13.2 (3.0) | 13.2 (3.2) |
| Activated partial thromboplastin time<br>(APTT) (seconds) | 29.6 (8.6) | 29.2 (8.8) |
| Sodium (mmol/L) | 137.0 (6.0) | 137.0 (6.0) |
| Potassium (mmol/L) | 4.1 (0.8) | 4.1 (0.7) |
| Total bilirubin (mg/dL) | 10.0 (7.0) | 10.0 (7.0) |
| Alanine aminotransferase (ALT)<br>(units/L) | 26.0 (27.0) | 25.0 (26.0) |
| Aspartate aminotransferase (AST)<br>(units/L) | 42.0 (41.0) | 48.0 (53.0) |
| Lactate dehydrogenase (Units/L) | 432.0 (330.2) | 416.5 (311.8) |
| Glucose (mmol/L) | 6.8 (3.1) | 6.8 (3.2) |
| Urea (mmol/L) | 7.1 (6.4) | 7.3 (6.8) |
| Creatinine (μmol/L) | 86.0 (53.0) | 86.0 (56.0) |
| Lactate (mmol/L) | 1.5 (1.0) | 1.5 (1.1) |
| C-reactive protein (CRP) (mg/dL) | 85.0 (121.0) | 78.0 (120.0) |

Values stated as median with IQR in parentheses for continuous variables, patient number with percentage in parentheses for categorical variables. Comorbidities were defined using the Charlson Comorbidity Index, with the addition of clinician-defined obesity. Information on missing data contained within Appendix 3.

### Model development

In total, 41 candidate predictor variables measured at hospital admission were identified for model creation (Figure 1; Appendix 2). Following the creation of a composite variable containing all seven individual comorbidities and the exclusion of 13 variables due to high levels of missing values (Appendix 3), 21 variables remained.

Generalised additive modelling (GAM) identified eight important predictors of mortality; age, sex, number of comorbidities, respiratory rate, peripheral oxygen saturation, Glasgow Coma Scale (GCS), urea, and C-reactive protein (CRP) (for variable selection process, see Appendix 4). Given the *a priori* need for a pragmatic score for use at the bedside, continuous variables were converted to factors with cut-points chosen using component smoothed functions (on linear predictor scale) from GAM model (Appendix 5).

On entering variables into a penalised logistic regression model (LASSO), all variables were retained within the final model (Appendix 6). Penalised regression coefficients were converted into a prognostic index using appropriate scaling (4C Mortality Score range 0-21 points; Table 2).

**Table 2.** Final 4C Mortality Score for in-hospital mortality in patients with covid-19. Prognostic index derived from penalised logistic regression (LASSO) model.

| Variable | 4C Mortality Score |  |
| --- | --- | --- |
| Age (years) | <50 |  |
|  | 50-59 | +2 |
|  | 60-69 | +4 |
|  | 70-79 | +6 |
|  | ≥80 | +7 |
| Sex at birth | Female |  |
|  | Male | +1 |
| Number of comorbidities* | 0 |  |
|  | 1 | +1 |
|  | ≥2 | +2 |
| Respiratory rate (breaths/minute) | <20 |  |
|  | 20-29 | +1 |
|  | ≥30 | +2 |
| Peripheral oxygen saturation on room air (%) | ≥92 |  |
|  | <92 | +2 |
| Glasgow Coma Scale | 15 |  |
|  | <15 | +2 |
| Urea (mmol/L) | ≤7 |  |
|  | 7-14 | +1 |
|  | >14 | +3 |
| CRP (mg/dL) | <50 |  |
|  | 50-99 | +1 |
|  | ≥100 | +2 |

\*Comorbidities were defined using the Charlson Comorbidity Index, with the addition of clinician-defined obesity

The 4C Mortality Score demonstrated good discrimination for in-patient death within the derivation cohort (Table 3) with performance that approached that of the XGBoost model. The 4C Mortality Score showed good calibration (Calibration slope = 1; Figure 2) across the range of risk and no adjustment to the model was required.

**Table 3.** Model discrimination in derivation cohort.

| Model | AUROC | 95% CI | Brier score |
| --- | --- | --- | --- |
| 4C Mortality Score | 0.79 | 0.78 to 0.79 | 0.174 |
| Machine learning comparison<br>(extreme gradient boosting [XGBoost]) | 0.81 | 0.79 to 0.82 | 0.179 |

AUROC, area under receiver operator curve; CI, confidence interval.

**Figure 2.**
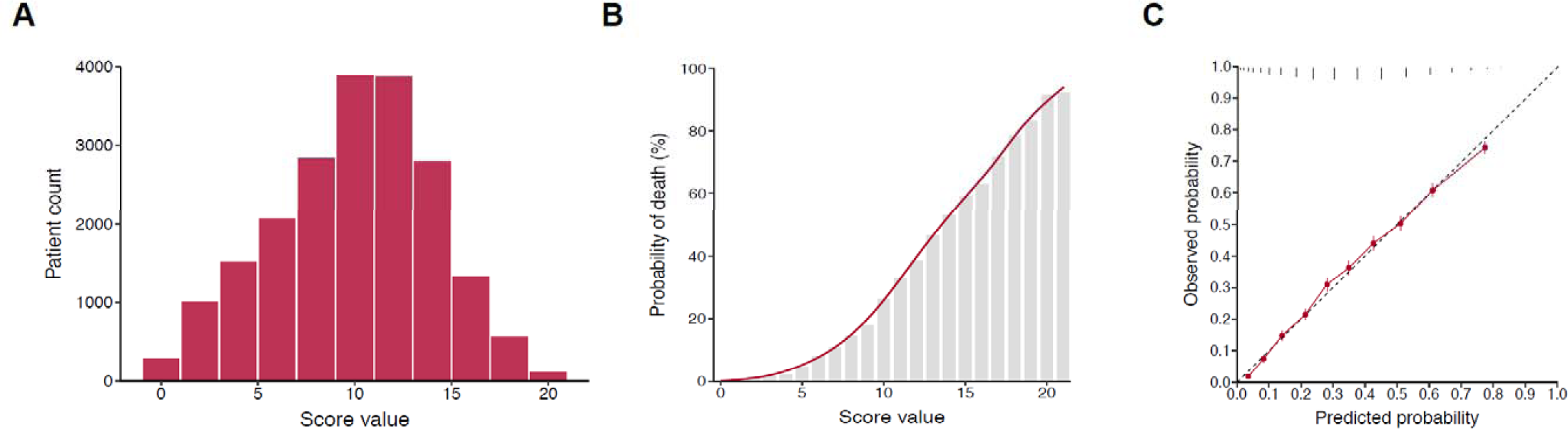
A, distribution of patients across range of 4C Mortality Score in derivation cohort. B, observed inpatient mortality across range of 4C Mortality Score in derivation cohort. C, predicted versus observed probability of inpatient mortality (calibration; red line) with distribution of patients across predicted probability (vertical black lines) for 4C Mortality Score within derivation cohort.

## Model validation

The validation cohort included data from 22 454 patients collected between 21^st^ May 2020 and 29^th^ June 2020 who had at least four weeks follow-up. The overall mortality was 28.6% (10 998 patients). The median age of patients in the cohort was 76 (interquartile range (IQR) 60-85) years. 12 202 (54.4%) were male and 16 930 patients (75.4%) had at least one comorbidity (Table 1). Missing data for predictor variables within the validation cohort are summarised in Appendix 7.

Discrimination of the 4C Mortality Score in the validation cohort was similar to that of the XGBoost model. Calibration was also found to be excellent in the validation cohort (Calibration slope = 1; Appendix 8), with a similar Brier score to the derivation cohort (0.174). The 4C Mortality Score demonstrated good performance in clinically relevant metrics, across a range of cut-offs (Table 4).

**Table 4.** Performance metrics of 4C Mortality Score to rule-out mortality (A) and rule-in mortality (B) at different cut-offs in validation cohort.

|  | Number of patients<br>at cut-off (%) | TP | TN | FP | FN | Sensitivity | Specificity | PPV | NPV | Mortality<br>(%) |
| --- | --- | --- | --- | --- | --- | --- | --- | --- | --- | --- |
| ≤2 | 512 (3.9) | 4310 | 510 | 8422 | 2 | 100.0 | 5.7 | 33.9 | 99.6 | 0.4 |
| ≤3 | 918 (6.9) | 4303 | 909 | 8023 | 9 | 99.8 | 10.2 | 34.9 | 99.0 | 1.0 |
| ≤4 | 1353 (10.2) | 4282 | 1323 | 7609 | 30 | 99.3 | 14.8 | 36.0 | 97.8 | 2.2 |
| ≤6 | 2417 (18.2) | 4196 | 2301 | 6631 | 116 | 97.3 | 25.8 | 38.8 | 95.2 | 4.8 |
| ≤8 | 3925 (29.6) | 4007 | 3620 | 5312 | 305 | 92.9 | 40.5 | 43.0 | 92.2 | 7.8 |

**B**
|  | Number of patients<br>at cut-off (%) | TP | TN | FP | FN | Sensitivity | Specificity | PPV | NPV | Mortality<br>(%) |
| --- | --- | --- | --- | --- | --- | --- | --- | --- | --- | --- |
| ≥9 | 9319 (70.4) | 4007 | 3620 | 5312 | 305 | 92.9 | 40.5 | 43.0 | 92.2 | 43.0 |
| ≥11 | 7109 (53.7) | 3489 | 5312 | 3620 | 823 | 80.9 | 59.5 | 49.1 | 86.6 | 49.1 |
| ≥13 | 4495 (33.9) | 2571 | 7008 | 1924 | 1741 | 59.6 | 78.5 | 57.2 | 80.1 | 57.2 |
| ≥15 | 2310 (17.4) | 1542 | 8164 | 768 | 2770 | 35.8 | 91.4 | 66.8 | 74.7 | 66.8 |

TP, true positive; TN, true negative; FP, false positive; FN, false negative; PPV, positive predictive value; NPV, negative predictive value.

Four risk groups were defined with corresponding mortality rates determined (Table 5): low risk (0-3 score; mortality rate 1.0%), intermediate risk (4-8 score; 9.8%), high risk (9-14 score; 35.2%), and very high risk (≥15 score; 66.8%). Performance metrics demonstrated a high sensitivity (99.8%) and negative predictive value (NPV; 99.0%) for the low-risk group, covering 6.9% of the cohort and a corresponding mortality rate of 1.0%. Patients in the intermediate risk group (score 4-8; n = 3007, 22.7%) had a mortality rate of 9.8% (NPV 90.2%). High-risk patients (score 9-14; n = 7009, 52.9%) had a 35.2% mortality (NPV 64.8), while patients scoring ≥15 (n = 2310, 17.4%) had a 66.8% mortality (positive predictive value 66.8%).

**Table 5.** Comparison of mortality rates for 4C Mortality Score risk groups across derivation and validation cohorts.

| Risk group | Derivation cohort |  | Validation cohort |  |
| --- | --- | --- | --- | --- |
|  | Number of<br>patients (%) | Mortality<br>(%) | Number of<br>patients (%) | Mortality<br>(%) |
| Low (0-3) | 1275 (6.5) | 1.5 | 918 (6.9) | 1.0 |
| Intermediate (4-8) | 4642 (23.8) | 9.9 | 3007 (22.7) | 9.8 |
| High (9-14) | 10 430 (53.4) | 38.1 | 7009 (52.9) | 35.2 |
| Very high (≥15) | 3175 (16.3) | 69.8 | 2310 (17.4) | 66.8 |
| Overall | 19522 | 34.2 | 13244 | 32.6 |

## Comparison with existing tools

A total of 15 risk stratification scores were identified through a systematic literature search ^6,20,27–39^ The 4C Mortality Score compared well against existing risk stratification scores in predicting inpatient mortality (Table 6). Risk stratification scores originally validated in patients with community-acquired pneumonia (n = 9) generally had higher discrimination for inpatient mortality in the validation cohort (e.g., A-DROP [AUROC 0.74; 95%CI 0.73-0.75], E-CURB65 [0.76; 0.74-0.79]) (Figure 3A) than those developed within covid-19 cohorts (n = 4: Surgisphere [0.63; 0.62-0.64], DL score [0.67; 0.66-0.68], COVID-GRAM [0.71; 0.68-0.74] and Xie score [0.73; 0.71-0.76]). Performance metrics for the 4C Mortality Score compared well against existing risk stratification scores at specified cut-offs (Appendix 10).

**Table 6.**
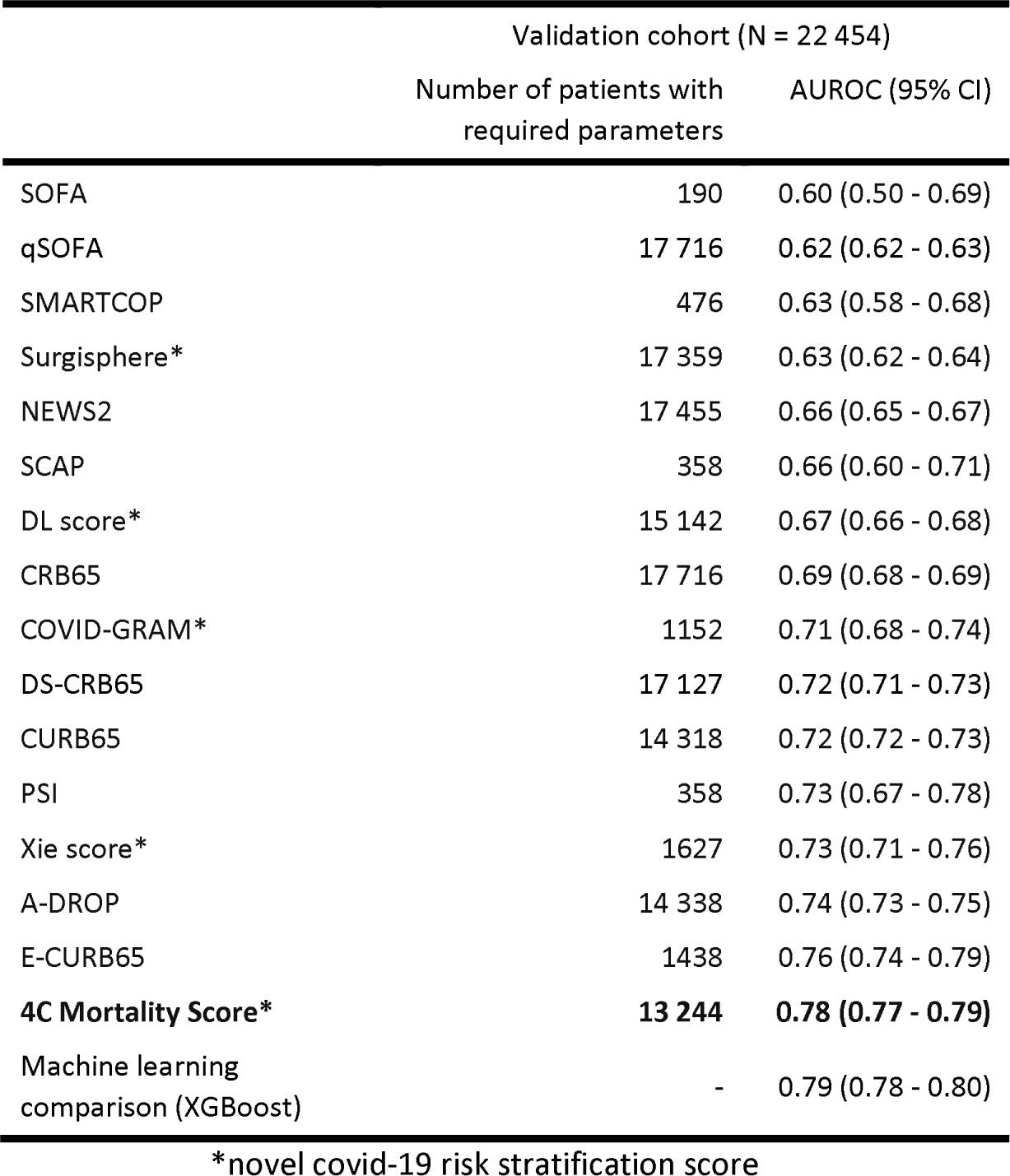
Discriminatory performance of risk stratification scores within validation cohort to predict inpatient mortality in patients hospitalised with covid-19. See appendix 10 for other metrics.

|  | Validation cohort (N = 22 454) |  |
| --- | --- | --- |
|  | Number of patients with required parameters | AUROC (95% CI) |
| SOFA | 190 | 0.60 (0.50 - 0.69) |
| qSOFA | 17 716 | 0.62 (0.62 - 0.63) |
| SMARTCOP | 476 | 0.63 (0.58 - 0.68) |
| Surgisphere* | 17 359 | 0.63 (0.62 - 0.64) |
| NEWS2 | 17 455 | 0.66 (0.65 - 0.67) |
| SCAP | 358 | 0.66 (0.60 - 0.71) |
| DL score* | 15 142 | 0.67 (0.66 - 0.68) |
| CRB65 | 17 716 | 0.69 (0.68 - 0.69) |
| COVID-GRAM* | 1152 | 0.71 (0.68 - 0.74) |
| DS-CRB65 | 17 127 | 0.72 (0.71 - 0.73) |
| CURB65 | 14 318 | 0.72 (0.72 - 0.73) |
| PSI | 358 | 0.73 (0.67 - 0.78) |
| Xie score* | 1627 | 0.73 (0.71 - 0.76) |
| A-DROP | 14 338 | 0.74 (0.73 - 0.75) |
| E-CURB65 | 1438 | 0.76 (0.74 - 0.79) |
| **4C Mortality Score\*** | **13 244** | **0.78 (0.77 - 0.79)** |
| Machine learning comparison (XGBoost) | - | 0.79 (0.78 - 0.80) |

\*novel covid-19 risk stratification score

**Figure 3.**
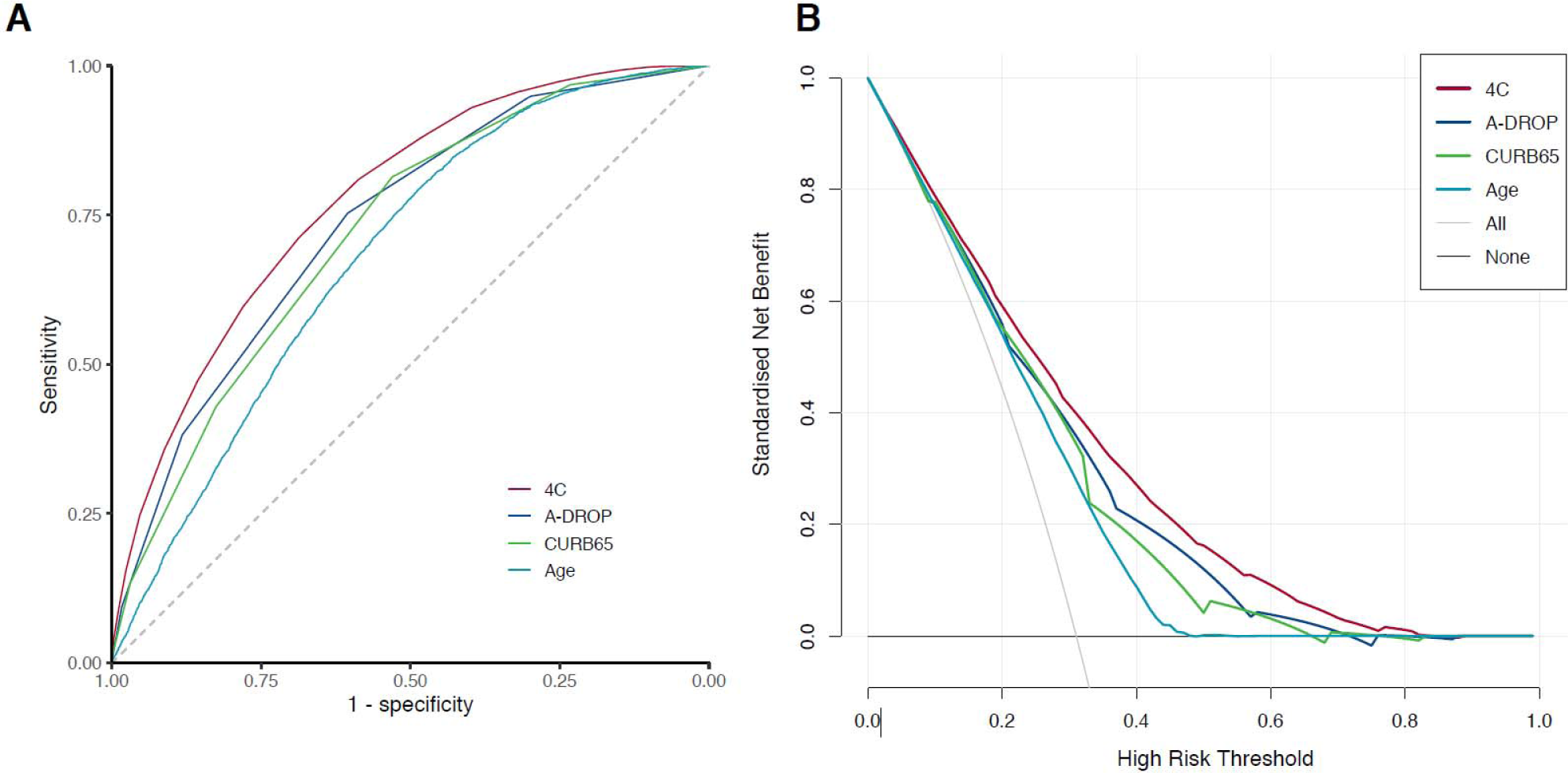
Receiver operator curves (ROC) (A) and decision curve analysis (B) for most discriminating three models applicable to >50% of validation population, together with age alone (spline). B, Lines are shown for standardised net benefit at different risk thresholds of treating no patients (black) and treating all patients (grey).

The number of patients in whom risk stratification scores could be applied differed due to certain variables not being available, either due to missingness or because they were not tested for/recorded in clinical practice. Seven scores could be applied to fewer than 2000 patients (<10%) in the validation cohort, due to the requirement for biomarkers or physiological parameters that were not routinely captured (e.g. lactate dehydrogenase [LDH]).

Decision curve analysis demonstrated that the 4C Mortality Score had better clinical utility across the range of threshold risks compared with the best performing existing scores applicable to >50% of the validation cohort (A-DROP and CURB65).

## Sensitivity analysis

Multiple imputation was performed in both the derivation and validation cohorts (predictor variables n = 41). Prognostic models derived using from the multiply imputed derivation cohort had poorer performance than models derived using complete case data. This was true whether performance was assessed in the complete case or multiply imputed validation datasets. As a sensitivity analysis, the final model was assessed in the multiply imputed datasets. Discriminatory performance demonstrated a small reduction (≤0.02 change in AUROC) (Appendix 11) across both derivation and validation cohorts.

After stratification of the validation cohort into two geographical cohorts (validation north and south; Appendix 12), discrimination remained similar for the 4C Mortality Score in both the North (AUROC 0.78, 95%CI 0.77-0.79) and South (0.77, 0.76-0.79) subsets (Appendix 13).

## Discussion

### Principal findings

We have developed and validated the eight-variable 4C Mortality Score for in-patient death in a UK prospective cohort of 57 146 patients hospitalised with covid-19. The 4C Mortality Score uses patient demographics, clinical observations, and blood parameters that are commonly available at the time of hospital admission and can accurately predict patients at a high risk of in-hospital death. It compared favourably to other models, including ‘best-in-class’ machine learning techniques and demonstrated consistent performance across the validation cohorts including good clinical utility in a decision curve analysis.

Model performance compared well against other generated models, with minimal loss in discrimination despite its pragmatic nature. A machine learning approach demonstrated a marginal improvement in discrimination, but at the cost of interpretability, the requirement for many more input variables, and the requirement for an app/website calculator limiting use at the bedside. The 4C Mortality Score demonstrated good applicability within the validation cohort (around 60% population) and consistency across all performance measures.

### Comparison with other studies

The 4C Mortality Score contains parameters reflecting patient demographics, comorbidity, physiology, and inflammation on hospital admission. It shares characteristics with existing prognostic scores for sepsis and community-acquired pneumonia, as well as for scores developed in covid-19 patients. Altered consciousness and high respiratory rate are included in most risk stratification scores for sepsis and community-acquired pneumonia,^19,20,27,28,31,32,35^ while elevated urea is also a common component.^19,20,27^ Increasing age is a strong predictor of inpatient mortality within our hospitalised covid-19 cohort and is commonly included in other existing covid-19 scores,^36,40,41^together with comorbidity^36,40,41^ and elevated CRP.^39,42^

Discriminatory performance of existing covid-19 scores applied to our cohort was lower than reported in derivation cohorts (DL score 0.74; COVID-GRAM 0.88; Xie score 0.98).^36,37,39^ The use of small inpatient cohorts from Wuhan, China for model development may have resulted in over-fitting,limiting generalisability in other cohorts.^37,39^ The Xie score demonstrated the highest discriminatory power (0.73), including age, lymphocyte count, LDH and peripheral oxygen saturations. However, we were only able to calculate this score for <10% of the validation cohort, as LDH is not routinely captured on admission in the UK.

Due to challenges of clinical data collection during an epidemic, missing data is common, with choice of predictors influenced by data availability.^39^ Complete case analysis often leads to exclusion of a substantial proportion of the original sample, subsequently leading to a loss of precision and power.^43^ However, the assessment of missing data on model performance in novel covid-19 risk stratification scores has been limited^36^ or unexplored^37,39^, potentially introducing bias and further limiting generalisability to other cohorts. We found discriminatory performance in both derivation and validation cohorts remained similar after the imputation of a wide range of variables (41), further supporting the validity of our findings.

The presence of comorbidities is handled differently in covid-19 prognostic scores, either included individually,^39,41^ given equal weight,^36^ or found to have no predictive effect.^37^ Recent evidence suggests an additive effect of comorbidity in covid-19 patients, with increasing number of comorbidities associated with poorer outcomes.^15^ In our cohort, the inclusion of individual comorbidities within the machine learning model conferred minimal additional discriminatory performance, supporting the inclusion of an overall comorbidity count.

### Strengths and limitations of this study

The ISARIC WHO CCP-UK study represents the largest prospectively collected covid-19 hospitalised patient cohort in the world and reflects the clinical data available in most economically developed healthcare settings. We developed a clinically applicable score with clear methodology and tested it against existing risk stratification scores in a large hospitalised patient cohort. It compared favourably to other prognostic tools, with good to excellent discrimination, calibration and performance characteristics.

The 4C Mortality Score has several methodological advantages over current covid-19 prognostic scores. The use of penalised regression methods, an event-to-variable ratio greater than 100 reducing the risk of model over-fitting,^44,45^ and the use of clinical parameters at first assessment increases the clinical applicability of the score and limits use of highly selective predictors prevalent in other risk stratification scores.^4,46^ In addition, the sensitivity analyses demonstrated that score performance was robust.

Our study has limitations. First, we were unable to evaluate the predictive performance of a number of existing scores that comprise a large number of parameters (for example APACHE II^47^), as well as several other covid-19 prognostic scores that include computed tomography findings or uncommonly measured biomarkers.^5^ In addition, several potentially relevant comorbidities, such as hypertension, previous myocardial infarction and stroke^15^ were not included in data collection. Their inclusion might have impacted upon or improved 4C Mortality Score performance and generalisability.

Second, a proportion of recruited patients had incomplete episodes and were thus excluded from the analysis. Selection bias is possible if patients with incomplete episodes, such as those with prolonged hospital admission, had a differential mortality risk to those with completed episodes. Nevertheless, the size of our patient cohort compares favourably to other datasets for model creation. Furthermore, the patient cohort on which the 4C Mortality Score was derived comprised hospitalised patients who were seriously ill (mortality rate of 30.5%) and were of advanced age (median age 74 years). Further external validation is required to determine whether the 4C Mortality Score is generalisable among younger patients and in settings outside the UK.

### Conclusions and policy implications

We have derived and validated an easy-to-use eight-variable risk stratification score that enables accurate stratification of hospitalised covid-19 patients by mortality risk at hospital presentation. Application within the validation cohorts demonstrated this tool may guide clinician decisions, including treatment escalation.

The key aim of clinical risk stratification scores is to support clinical management decisions. Three risk classes were identified and demonstrated similar adverse outcome rates across the validation cohort. Patients with a 4C Mortality Score falling within the low-risk groups (mortality rate 1%) might be suitable for management in the community, while those within the intermediate-risk group were at lower risk of mortality (mortality rate 10%; 23% of the cohort) and may be suitable for ward-level monitoring. Similar mortality rates have been identified as an appropriate cut-off in pneumonia risk stratification scores (CURB-65 and PSI).^19,20^ Meanwhile patients with a score ≥9 were at high risk of death (43%), which may prompt aggressive treatment, including the commencement of steroids,^48^ and early escalation to critical care if appropriate.

## Data Availability

We welcome applications for data and material access via our Independent Data And Material Access Committee (https://isaric4c.net).

https://isaric4c.net/sample_access

## End matter

### What is already known on this topic

- There is a lack of robust, validated clinical prediction tools to identify patients with covid-19 who are at the highest risk of mortality
- Given uncertainty about how to stratify covid-19 patients, there is considerable interest in risk stratification scores to support frontline clinical decision making
- Available risk stratification tools however suffer from a high risk of bias, small sample size resulting in uncertainty, poor reporting and lack of formal validation

### What this study adds

- The majority of existing covid-19 risk stratification tools performed poorly in our cohort – caution should be applied when using novel tools based on small patient populations to in-hospital cohorts with covid-19
- In contrast, our 4C (Coronavirus Clinical Characterisation Consortium) score is an easy-to-use and valid prediction tool for inpatient mortality, accurately categorising patients as being at low, intermediate, high, or very high-risk of death
- This pragmatic and clinically applicable score outperformed other risk stratification tools and had similar performance to more complex models

### Statements

The study protocol is available at http://isaric4c.net/protocols; study registry https://www.isrctn.com/ISRCTN66726260.

The Corresponding Author has the right to grant on behalf of all authors and does grant on behalf of all authors, a worldwide licence (http://www.bmj.com/sites/default/files/BMJ%20Author%20Licence%20March%202013.doc) to the Publishers and its licensees in perpetuity, in all forms, formats and media (whether known now or created in the future), to i) publish, reproduce, distribute, display and store the Contribution, ii) translate the Contribution into other languages, create adaptations, reprints, include within collections and create summaries, extracts and/or, abstracts of the Contribution and convert or allow conversion into any format including without limitation audio, iii) create any other derivative work(s) based in whole or part on the on the Contribution, iv) to exploit all subsidiary rights to exploit all subsidiary rights that currently exist or as may exist in the future in the Contribution, v) the inclusion of electronic links from the Contribution to third party material where-ever it may be located; and, vi) licence any third party to do any or all of the above.

## Acknowledgments

This work uses data provided by patients and collected by the NHS as part of their care and support #DataSavesLives. We are extremely grateful to the 2,648 frontline NHS clinical and research staff and volunteer medical students, who collected this data in challenging circumstances; and the generosity of the participants and their families for their individual contributions in these difficult times. We also acknowledge the support of Jeremy J Farrar, Nahoko Shindo, Devika Dixit, Nipunie Rajapakse, Piero Olliaro, Lyndsey Castle, Martha Buckley, Debbie Malden, Katherine Newell, Kwame O’Neill, Emmanuelle Denis, Claire Petersen, Scott Mullaney, Sue MacFarlane, Chris Jones, Nicole Maziere, Katie Bullock, Emily Cass, William Reynolds, Milton Ashworth, Ben Catterall, Louise Cooper, Terry Foster, Paul Matthew Ridley, Anthony Evans, Catherine Hartley, Chris Dunn, Debby Sales, Diane Latawiec, Erwan Trochu, Eve Wilcock, Innocent Gerald Asiimwe, Isabel Garcia-Dorival, J. Eunice Zhang, Jack Pilgrim, Jane A Armstrong, Jordan J. Clark, Jordan Thomas, Katharine King, Katie Alexandra Ahmed, Krishanthi S Subramaniam, Lauren Lett, Laurence McEvoy, Libby van Tonder, Lucia Alicia Livoti, Nahida S Miah, Rebecca K. Shears, Rebecca Louise Jensen, Rebekah Penrice-Randal, Robyn Kiy, Samantha Leanne Barlow, Shadia Khandaker, Soeren Metelmann, Tessa Prince, Trevor R Jones, Benjamin Brennan, Agnieska Szemiel, Siddharth Bakshi, Daniella Lefteri, Maria Mancini, Julien Martinez, Angela Elliott, Joyce Mitchell, John McLauchlan, Aislynn Taggart, Oslem Dincarslan, Annette Lake, Claire Petersen, and Scott Mullaney.

## ISARIC Coronavirus Clinical Characterisation Consortium (ISARIC4C) Investigators

Consortium Lead Investigator J Kenneth Baillie, Chief Investigator Malcolm G Semple, Co-Lead Investigator Peter JM Openshaw. ISARIC Clinical Coordinator Gail Carson. Co-Investigators: Beatrice Alex, Benjamin Bach, Wendy S Barclay, Debby Bogaert, Meera Chand, Graham S Cooke, Annemarie B Docherty, Jake Dunning, Ana da Silva Filipe, Tom Fletcher, Christopher A Green, Ewen M Harrison, Julian A Hiscox, Antonia Ying Wai Ho, Peter W Horby, Samreen Ijaz, Saye Khoo, Paul Klenerman, Andrew Law, Wei Shen Lim, Alexander, J Mentzer, Laura Merson, Alison M Meynert, Mahdad Noursadeghi, Shona C Moore, Massimo Palmarini, William A Paxton, Georgios Pollakis, Nicholas Price, Andrew Rambaut, David L Robertson, Clark D Russell, Vanessa Sancho-Shimizu, Janet T Scott, Louise Sigfrid, Tom Solomon, Shiranee Sriskandan, David Stuart, Charlotte Summers, Richard S Tedder, Emma C Thomson, Ryan S Thwaites, Lance CW Turtle, Maria Zambon. Project Managers Hayley Hardwick, Chloe Donohue, Jane Ewins, Wilna Oosthuyzen, Fiona Griffiths. Data Analysts: Lisa Norman, Riinu Pius, Tom M Drake, Cameron J Fairfield, Stephen Knight, Kenneth A Mclean, Derek Murphy, Catherine A Shaw. Data and Information System Manager: Jo Dalton, Michelle Girvan, Egle Saviciute, Stephanie Roberts Janet Harrison, Laura Marsh, Marie Connor. Data integration and presentation: Gary Leeming, Andrew Law, Ross Hendry. Material Management: William Greenhalf, Victoria Shaw, Sarah McDonald. Outbreak Laboratory Volunteers: Katie A. Ahmed, Jane A Armstrong, Milton Ashworth, Innocent G Asiimwe, Siddharth Bakshi, Samantha L Barlow, Laura Booth, Benjamin Brennan, Katie Bullock, Benjamin WA Catterall, Jordan J Clark, Emily A Clarke, Sarah Cole, Louise Cooper, Helen Cox, Christopher Davis, Oslem Dincarslan, Chris Dunn, Philip Dyer, Angela Elliott, Anthony Evans, Lewis WS Fisher, Terry Foster, Isabel Garcia-Dorival, Willliam Greenhalf, Philip Gunning, Catherine Hartley, Antonia Ho, Rebecca L Jensen, Christopher B Jones, Trevor R Jones, Shadia Khandaker, Katharine King, Robyn T. Kiy, Chrysa Koukorava, Annette Lake, Suzannah Lant, Diane Latawiec, L Lavelle-Langham, Daniella Lefteri, Lauren Lett, Lucia A Livoti, Maria Mancini, Sarah McDonald, Laurence McEvoy, John McLauchlan, Soeren Metelmann, Nahida S Miah, Joanna Middleton, Joyce Mitchell, Shona C Moore, Ellen G Murphy, Rebekah Penrice-Randal, Jack Pilgrim, Tessa Prince, Will Reynolds, P. Matthew Ridley, Debby Sales, Victoria E Shaw, Rebecca K Shears, Benjamin Small, Krishanthi S Subramaniam, Agnieska Szemiel, Aislynn Taggart, Jolanta Tanianis, Jordan Thomas, Erwan Trochu, Libby van Tonder, Eve Wilcock, J. Eunice Zhang. Local Principal Investigators: Kayode Adeniji, Daniel Agranoff, Ken Agwuh, Dhiraj Ail, Ana Alegria, Brian Angus, Abdul Ashish, Dougal Atkinson, Shahedal Bari, Gavin Barlow, Stella Barnass, Nicholas Barrett, Christopher Bassford, David Baxter, Michael Beadsworth, Jolanta Bernatoniene, John Berridge, Nicola Best, Pieter Bothma, David Brealey, Robin Brittain-Long, Naomi Bulteel, Tom Burden, Andrew Burtenshaw, Vikki Caruth, David Chadwick, Duncan Chambler, Nigel Chee, Jenny Child, Srikanth Chukkambotla, Tom Clark, Paul Collini, Catherine Cosgrove, Jason Cupitt, Maria-Teresa Cutino-Moguel, Paul Dark, Chris Dawson, Samir Dervisevic, Phil Donnison, Sam Douthwaite, Ingrid DuRand, Ahilanadan Dushianthan, Tristan Dyer, Cariad Evans, Chi Eziefula, Chrisopher Fegan, Adam Finn, Duncan Fullerton, Sanjeev Garg, Sanjeev Garg, Atul Garg, Jo Godden, Arthur Goldsmith, Clive Graham, Elaine Hardy, Stuart Hartshorn, Daniel Harvey, Peter Havalda, Daniel B Hawcutt, Maria Hobrok, Luke Hodgson, Anita Holme, Anil Hormis, Michael Jacobs, Susan Jain, Paul Jennings, Agilan Kaliappan, Vidya Kasipandian, Stephen Kegg, Michael Kelsey, Jason Kendall, Caroline Kerrison, Ian Kerslake, Oliver Koch, Gouri Koduri, George Koshy, Shondipon Laha, Susan Larkin, Tamas Leiner, Patrick Lillie, James Limb, Vanessa Linnett, Jeff Little, Michael MacMahon, Emily MacNaughton, Ravish Mankregod, Huw Masson, Elijah Matovu, Katherine McCullough, Ruth McEwen, Manjula Meda, Gary Mills, Jane Minton, Mariyam Mirfenderesky, Kavya Mohandas, Quen Mok, James Moon, Elinoor Moore, Patrick Morgan, Craig Morris, Katherine Mortimore, Samuel Moses, Mbiye Mpenge, Rohinton Mulla, Michael Murphy, Megan Nagel, Thapas Nagarajan, Mark Nelson, Igor Otahal, Mark Pais, Selva Panchatsharam, Hassan Paraiso, Brij Patel, Justin Pepperell, Mark Peters, Mandeep Phull, Stefania Pintus, Jagtur Singh Pooni, Frank Post, David Price, Rachel Prout, Nikolas Rae, Henrik Reschreiter, Tim Reynolds, Neil Richardson, Mark Roberts, Devender Roberts, Alistair Rose, Guy Rousseau, Brendan Ryan, Taranprit Saluja, Aarti Shah, Prad Shanmuga, Anil Sharma, Anna Shawcross, Jeremy Sizer, Richard Smith, Catherine Snelson, Nick Spittle, Nikki Staines, Tom Stambach, Richard Stewart, Pradeep Subudhi, Tamas Szakmany, Kate Tatham, Jo Thomas, Chris Thompson, Robert Thompson, Ascanio Tridente, Darell Tupper - Carey, Mary Twagira, Andrew Ustianowski, Nick Vallotton, Lisa Vincent-Smith, Shico Visuvanathan, Alan Vuylsteke, Sam Waddy, Rachel Wake, Andrew Walden, Ingeborg Welters, Tony Whitehouse, Paul Whittaker, Ashley Whittington, Meme Wijesinghe, Martin Williams, Lawrence Wilson, Sarah Wilson, Stephen Winchester, Martin Wiselka, Adam Wolverson, Daniel G Wooton, Andrew Workman, Bryan Yates, Peter Young.

## Contributions

MG Semple is guarantor and corresponding author for this work, and attests that all listed authors meet authorship criteria and that no others meeting the criteria have been omitted.

### Contributor role taxonomy (CRediT)

Conceptualisation SRK, AH, RP, GC, JD, PWH, LM, JSN-V-T, CS, PJMO, JKB, MGS, ABD, EMH, Data curation: RP, SH, KAH, CJ, KAM, LM, LN, CDR, CAS, MGS, ABD, EMH. Formal analysis: SRK, AH, RP, TMD, CJF, KAM, MGP, LT, EMH. Funding acquisition: PWH, JSN-V-T, TS, PJMO, JKB, MGS, ABD. Investigation: SRK, AH, CDR, OVS, LT, PJMO, JKB, MGS, ABD, EMH. Methodology: SRK, AH, RP, IB, KAM, CAS, AS, CS, ABD, EMH. Project administration: SH, HEH, CJ, LM, LN, CDR, CAS, MGS. Resources: CG, PWH, LM, TS, MGS, EMH. Software: SRK, RP, KAM, OVS. Supervision: SRK, RP, CG, HEH, PWH, PLO, PJMO, JKB, MGS, EMH. Visualisation: SRK, RP, MGP, EMH. Writing-original draft: SRK, EMH. Writing-review and editing: SRK, RG, AH, RP, IB, GC, TMD, JD, CJF, LM, MN, JSN-V-T, PLO, MGP, CDR, AS, CS, OVS, LT, PJMO, JKB, MGS, ABD, EMH.

## Ethical approval

Ethical approval was given by the South Central – Oxford C Research Ethics Committee in England (Ref: 13/SC/0149), and by the Scotland A Research Ethics Committee (Ref: 20/SS/0028). The study was registered at https://www.isrctn.com/ISRCTN66726260.

## Data sharing

### Dissemination to participants and related patient and public communities

ISARIC4C has a public facing website isaric4c.net and twitter account @CCPUKstudy. We are engaging with print and internet press, television, radio, news, and documentary programme makers. We will explore distribution of findings with The Asthma UK and British Lung Foundation Partnership and take advice from NIHR Involve and GenerationR Alliance Young People’s Advisory Groups.

